# Emotions in the time of COVID-19: A sentiment analysis of tweets during the nationwide lockdown in India

**DOI:** 10.1101/2022.06.19.22276620

**Authors:** Rizwan Suliankatchi Abdulkader, Kathiresan Jeyashree, Deneshkumar Venugopal, K Senthamarai Kannan, Manickam Ponnaiah, Manoj Murhekar

## Abstract

**Background:** COVID-19 pandemic is unprecedented in terms of burden, nature and quantum of control measures and public reactions. We report trends in public emotions and sentiments before and during the nation-wide lockdown implemented since 25^th^ March 2020 in India.

**Methods:** We collected a sample of tweets containing the keywords ‘coronavirus’ or ‘COVID-19’ published between 12^th^ March and 14^th^ April in India. After pre-processing, the tweets were subjected to sentiment analysis using natural language processing algorithms.

**Results:** Our analysis of 226170 tweets revealed a positive public sentiment (mean sentiment score=0.25). Tweets expressing a given sentiment showed significant (p<0.001) waning of negativity; negative tweets decreased (39.3% to 35.9%) and positive tweets increased (49.8% to 51.8%). Trust (0.85 words/tweet/day) and fear (0.66 words/tweet/day) were the dominant positive and negative emotions, respectively.

**Conclusions:** Positive sentiments dominated during the COVID-19 lockdown in India. A surveillance system monitoring public sentiments on public health interventions for COVID-19 should be established.

## 1. Introduction

Public health responses to crisis situations such as pandemics are increasingly becoming dependent on online channels of communication. In the context of the ongoing COVID-19 pandemic, a large number of official communications, awareness messages, announcements of interventions and public health surveillance and action taken reports are communicated to the public via social media. These media also provide the space for the public to register their response, comments and opinions on the same and express themselves. Social media holds a mirror to these expressions and when analysed properly can provide valuable lessons for public health decision making. Recently, several computationally intensive methods have been developed to gain insights from social media via natural language processing and machine learning methods.(Kumar and Sebastian 2012)

Understanding the emotions around a particular illness is key to developing appropriate public health strategies to combat the illness.(Braunack-Mayer et al. 2010) What the public feel about a given situation and their opinions and emotions about a public health intervention will reflect in their actions.(Kim and Niederdeppe 2013) Their acceptance of and compliance with the strategy is not only vital for its optimal implementation but also cause the least public unrest and discomfort. Public risk perceptions and behaviors have been shown to follow media logic, rather than epidemiological logic.(Reintjes et al. 2016) Thus, the need for public health policy makers to keep track of public sentiments and factoring these into drafting acceptable public health strategies cannot be overemphasised.

There have been very few efforts in the field of health to understand the public sentiment in response to certain public health strategies. Loft et al, in their analysis of Facebook (FB) posts, reported that following the introduction of a HPV vaccination campaign, the FB page created for the purpose successfully reached and engaged FB users.(Loft et al. 2020) Gabarron et al while analysing the sentiments around Diabetes on Twitter concluded that such understanding is essential for promoting a positive and constructive attitude among people that read and discuss about the illness on social media.(Gabarron et al. 2019) Kang GJ et al found that sentiment analysis helped them in understanding the scope and variability of attitudes and beliefs toward vaccines.(Kang et al. 2017)

Public sentiments during a pandemic can span a wide range. The COVID-19 pandemic is unprecedented not just in terms of the disease burden but also in the magnitude of the public health response. Countries across the world have implemented very stringent measures restricting public movement and social interaction to curb this pandemic. Liu et al who report the results of their topic modelling exercise on news articles about COVID-19 in China recommend that future research should address the mass media’s actual impact on readers during the COVID-19 crisis through sentiment analysis.(Liu et al. 2020)

India is one of the countries currently facing a burgeoning load of COVID cases and deaths. In addition to the already existing ban on international travel and rigorous surveillance and contact tracing, the country implemented stringent control measures like closure of educational institutions, suspending public transport systems and a nationwide lockdown allowing only essential services to continue. The first phase of this lock down for 21 days was rolled out by the Government of India on the 25^th^ of March 2020.(Ministry of Home Affairs, n.d.) In this paper, we aim to describe the trends in public emotions and sentiments in a representative sample of tweets about COVID-19 and detect changes in response to the announcement of the complete lockdown in India.

## 2. Methods

### 2.1. Data collection

We collected tweets containing the keywords ‘coronavirus’ or ‘COVID-19’ published between 12^th^ March 2020 and 14^th^ April 2020 in the Twitter official website. The complete and mandatory lockdown in India began on 25^th^ March 2020 for a period of 21 days. This was preceded by a one day voluntary lockdown on Sunday, the 21^st^ of March 2020. The study period was chosen to encompass these milestone events. Tweets posted from within a 200 mile radius of five major cities of India (Bhopal, Chennai, Delhi, Kolkata and Mumbai) were extracted. This ensured a reasonable geographical representation of the entire nation. Tweets from neighbouring countries (Bangladesh) were excluded by using the ‘NOT’ operator in the search query. Completeness of the data extraction was verified by checking that the timestamp on the tweets spanned every hour on all included days. Therefore, tweets representing every hour of every day of the study period had been extracted. The standard Twitter application program interface (API) which provides the tweets and their metadata for predefined search parameters was used to collect data via the ‘Tweepy’ library in Python v. 3.6.10. We extracted tweet ID, timestamp, text of the tweet, emojis used and primary tweet language for every tweet. No limit was set on the number of tweets to be extracted and the number retrieved was managed by the inbuilt limitations of the standard Twitter API.

### 2.2. Data pre-processing

The initial steps in pre-processing involved removal of duplicate tweets using tweet ID as the unique identifier and non-English tweets using the tweet language variable. Further steps involved cleaning the text of the tweets by removing twitter handles, user mentions, hashtags, hyperlinks, links to images, punctuation marks, line breaks and extra white spaces, converting to lower case and changing the Universal Time Coordinated (UTC) timestamp to Indian Standard Time (IST) timestamp. Wherever emojis were encountered in tweets, the text description of the respective emojis were extracted and added to the main tweet text. For example, ☺ emoji was converted into the descriptive text ‘smiling face’ and added to the text.

### 2.3. Statistical analysis

The cleaned dataset containing the texts of the tweets was subjected to sentiment analysis using the ‘syuzhet’ package in R v.3.6.3.(“Introduction to the Syuzhet Package” n.d.) The ‘syuzhet’ lexicon was developed at the Nebraska Literary Lab and it attempts to reveal the latent narrative structure and emotional shifts between conflict and conflict resolution. This lexicon was based on a collection of 165,000 human coded sentences from a corpus of contemporary novels. It can only work with languages that use Latin character sets, making it unsuitable for most Indian languages and hence the need to exclude non-English tweets.(Phani, Lahiri, and Biswas 2016)

The first step in sentiment analysis was to obtain the sentiment score for each tweet. The sentiment score is a numeric vector where a negative number indicates a negative sentiment, zero indicates neutral sentiment and a positive number indicates a positive sentiment and the absolute value indicates the magnitude of the sentiment. The proportion of tweets representing each sentiment was compared for the period before and after the lockdown was compared using chi square test. We calculated the mean sentiment score for each day in each of the five cities and for the country as a whole, creating a daily time series of sentiment scores.

The second step was to count the number of words in each tweet that mapped to one of the eight emotions (anger, fear, anticipation, trust, surprise, sadness, joy, and disgust) and two sentiments (positive and negative). This was done based on the NRC Word-Emotion Association Lexicon.(“NRC Emotion Lexicon” n.d.) This lexicon is a list of English words that have been manually and reliably coded and associated with one or more of the above mentioned predefined basic emotions and sentiments. We calculated the mean number of words representing the eight emotions and two sentiments per tweet per day.

We performed change point analysis to assess how the trends in sentiments and emotions changed over time. Change points were described in terms of the ‘days since lockdown’ and whether an increase or decrease was observed at each change point.

Finally, we created a word cloud representing the 100 words that most commonly appeared in the tweets, categorized for the periods before and during the lockdown period. This was achieved using the text mining package ‘tm’ in R v.3.6.3.(Feinerer 2019) The ‘tm’ package provides for the preprocessing of tweets in terms of removing English stop words, user defined search keywords, punctuations and numbers and the creation of a sparse matrix called the term document matrix. This term document matrix formed the basis for the creation of the word cloud.

### 2.4. Data privacy and ethical issues

The tweets analyzed in this study were publicly available on the Internet. Users agree to the terms and conditions of using the Twitter service at the time of registration and also agree to regular updates. It is understood that an implied consent is given by the users for their data to be used for research purposes.(“Twitter Terms of Service” n.d.; Sloan et al. 2013) Our study is a secondary analysis of the texts contained in those tweets and entailed no harm to the users of Twitter or any other party. Our results only present summaries in the form of means and percentages and do not mention any particular tweet or user name or twitter handle, protecting the anonymity and privacy of the users. The analysis carried out here is based on an automated and reproducible process called natural language processing and does not involve a human reading or interpreting individual tweets.

## 3. Results

### 3.1. Sentiments

The sentiment analysis of 2,26,170 tweets from five major cities of India between 12.3.20 to 14.4.20 revealed that the mean sentiment score was 0.25. The mean sentiment score before the implementation of the lockdown was 0.16 and during the lockdown it was 0.26. The mean number of positive and negative words per tweet per day were 1.35 and 0.98, respectively. While positive words increased from 1.33 to 1.35 during the lockdown, the negative words reduced from 1.11 to 0.96. (Table 1, Figures 1,2) Before the lockdown, the percentage of positive, neutral and negative tweets were 49.8%, 10.9% and 39.3%, respectively. During the lockdown period, the percentage of positive tweets increased to 51.8% and so did the neutral tweets to 12.3% whereas the negative tweets decreased to 35.9% (chi square = 150.5, p value <0.001). Hence both before and during the intervention, positively oriented tweets were predominant.(Table 1)

**Table 1.**
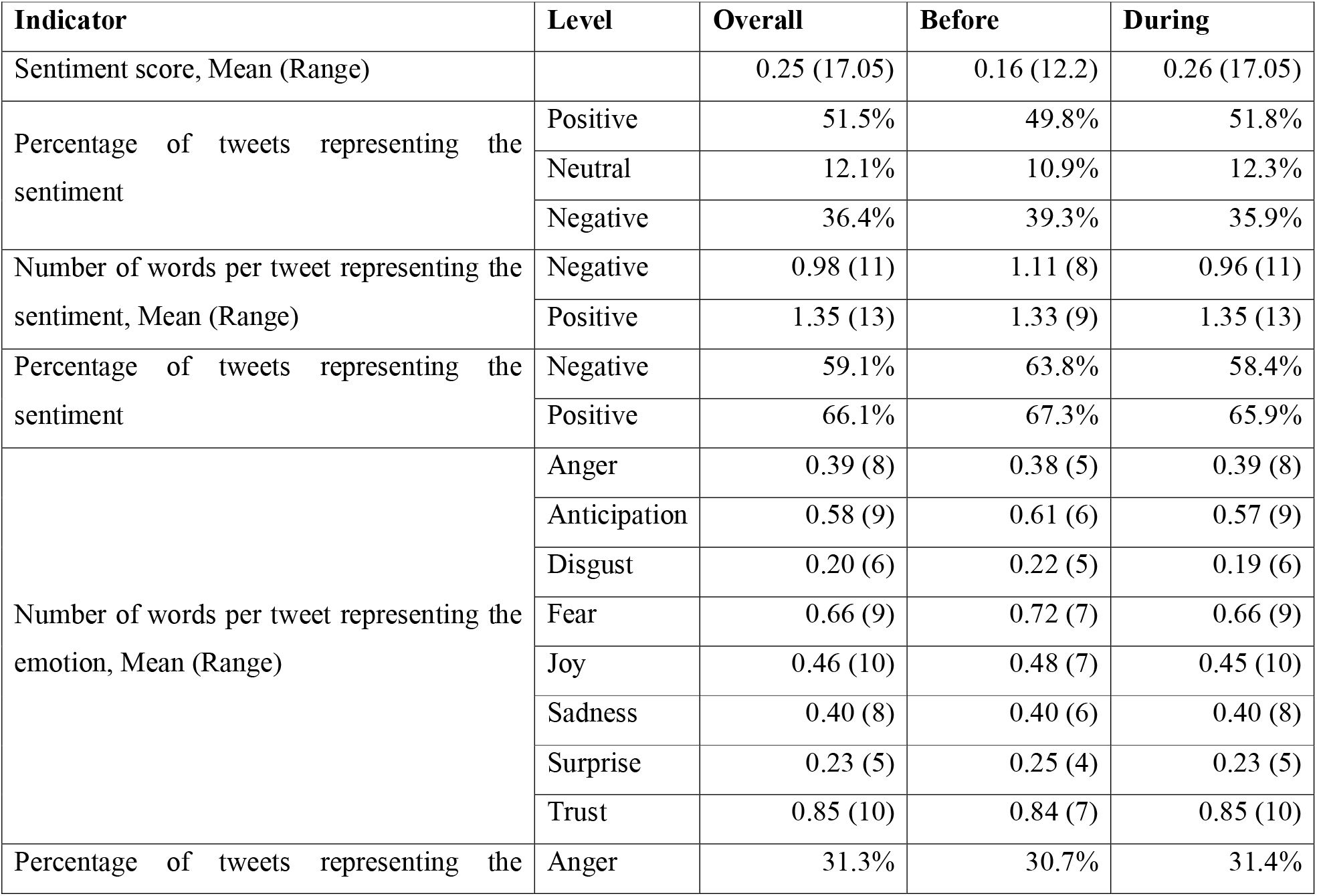
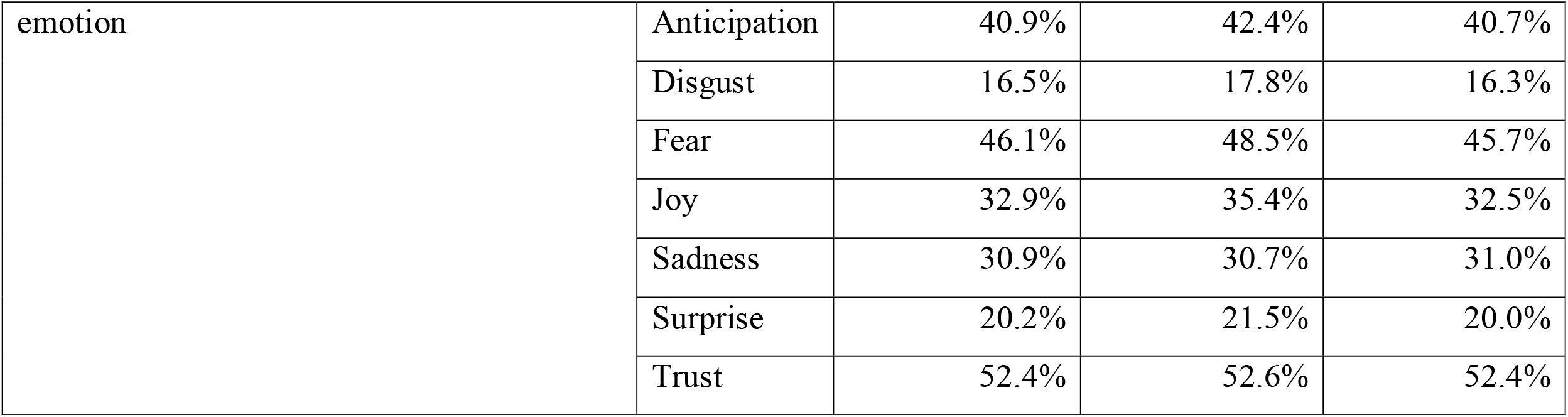
Public sentiments and emotions about COVID-19 before (12.3.20 to 24.3.20) and during (25.3.20 to 14.4.20) the first phase of lockdown in India.

| Indicator | Level | Overall | Before | During |
| --- | --- | --- | --- | --- |
| Sentiment score, Mean (Range) |  | 0.25 (17.05) | 0.16 (12.2) | 0.26 (17.05) |
| Percentage of tweets representing the sentiment | Positive | 51.5% | 49.8% | 51.8% |
|  | Neutral | 12.1% | 10.9% | 12.3% |
|  | Negative | 36.4% | 39.3% | 35.9% |
| Number of words per tweet representing the sentiment, Mean (Range) | Negative | 0.98 (11) | 1.11 (8) | 0.96 (11) |
|  | Positive | 1.35 (13) | 1.33 (9) | 1.35 (13) |
| Percentage of tweets representing the sentiment | Negative | 59.1% | 63.8% | 58.4% |
|  | Positive | 66.1% | 67.3% | 65.9% |
| Number of words per tweet representing the emotion, Mean (Range) | Anger | 0.39 (8) | 0.38 (5) | 0.39 (8) |
|  | Anticipation | 0.58 (9) | 0.61 (6) | 0.57 (9) |
|  | Disgust | 0.20 (6) | 0.22 (5) | 0.19 (6) |
|  | Fear | 0.66 (9) | 0.72 (7) | 0.66 (9) |
|  | Joy | 0.46 (10) | 0.48 (7) | 0.45 (10) |
|  | Sadness | 0.40 (8) | 0.40 (6) | 0.40 (8) |
|  | Surprise | 0.23 (5) | 0.25 (4) | 0.23 (5) |
|  | Trust | 0.85 (10) | 0.84 (7) | 0.85 (10) |
| Percentage of tweets representing the | Anger | 31.3% | 30.7% | 31.4% |
| emotion | Anticipation | 40.9% | 42.4% | 40.7% |
|  | Disgust | 16.5% | 17.8% | 16.3% |
|  | Fear | 46.1% | 48.5% | 45.7% |
|  | Joy | 32.9% | 35.4% | 32.5% |
|  | Sadness | 30.9% | 30.7% | 31.0% |
|  | Surprise | 20.2% | 21.5% | 20.0% |
|  | Trust | 52.4% | 52.6% | 52.4% |

**Figure 1.**
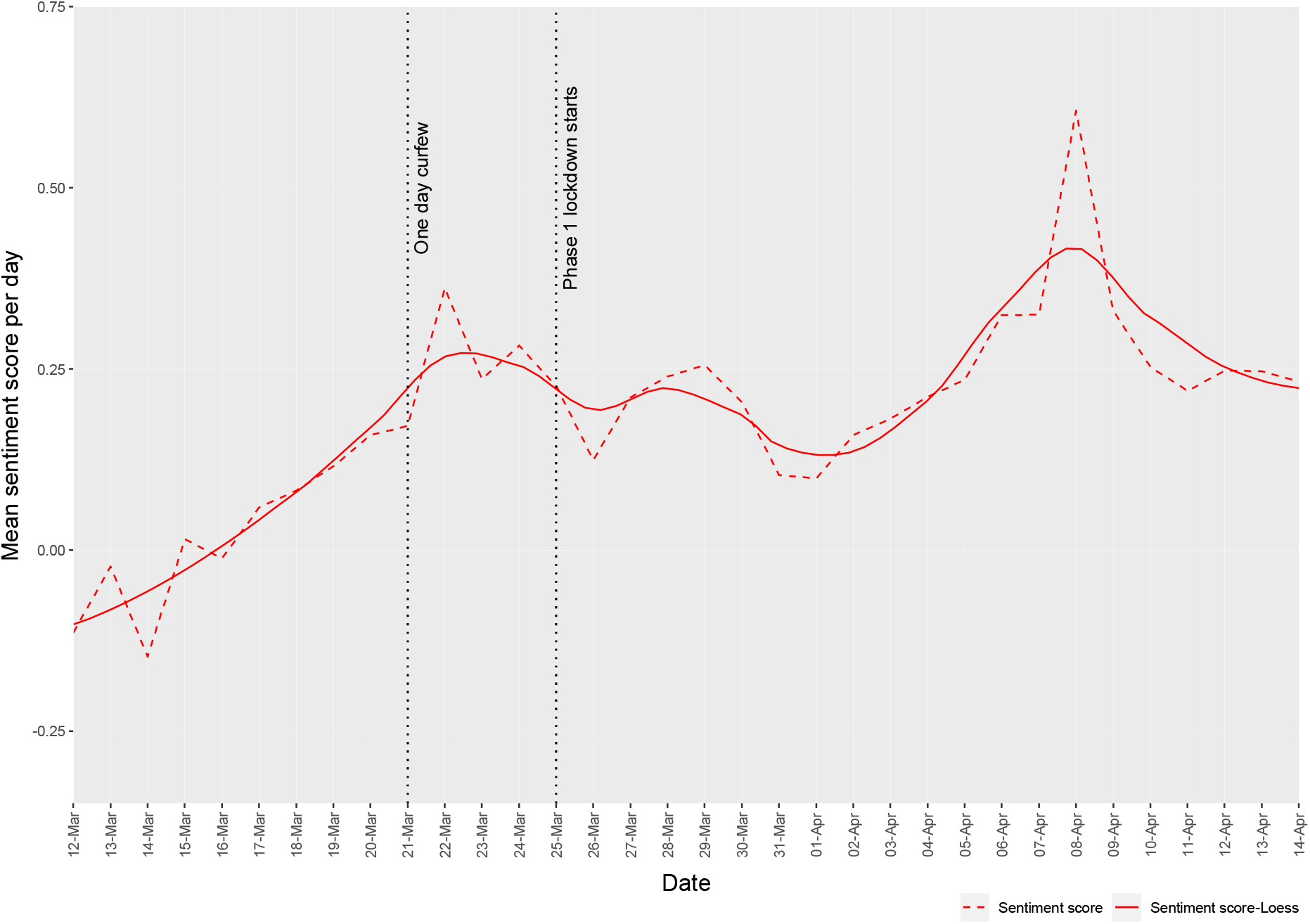
Trends in daily mean sentiment score before (12.3.20 to 24.3.20) and during (25.3.20 to 14.4.20) the first phase of lockdown in India.

**Figure 2.**
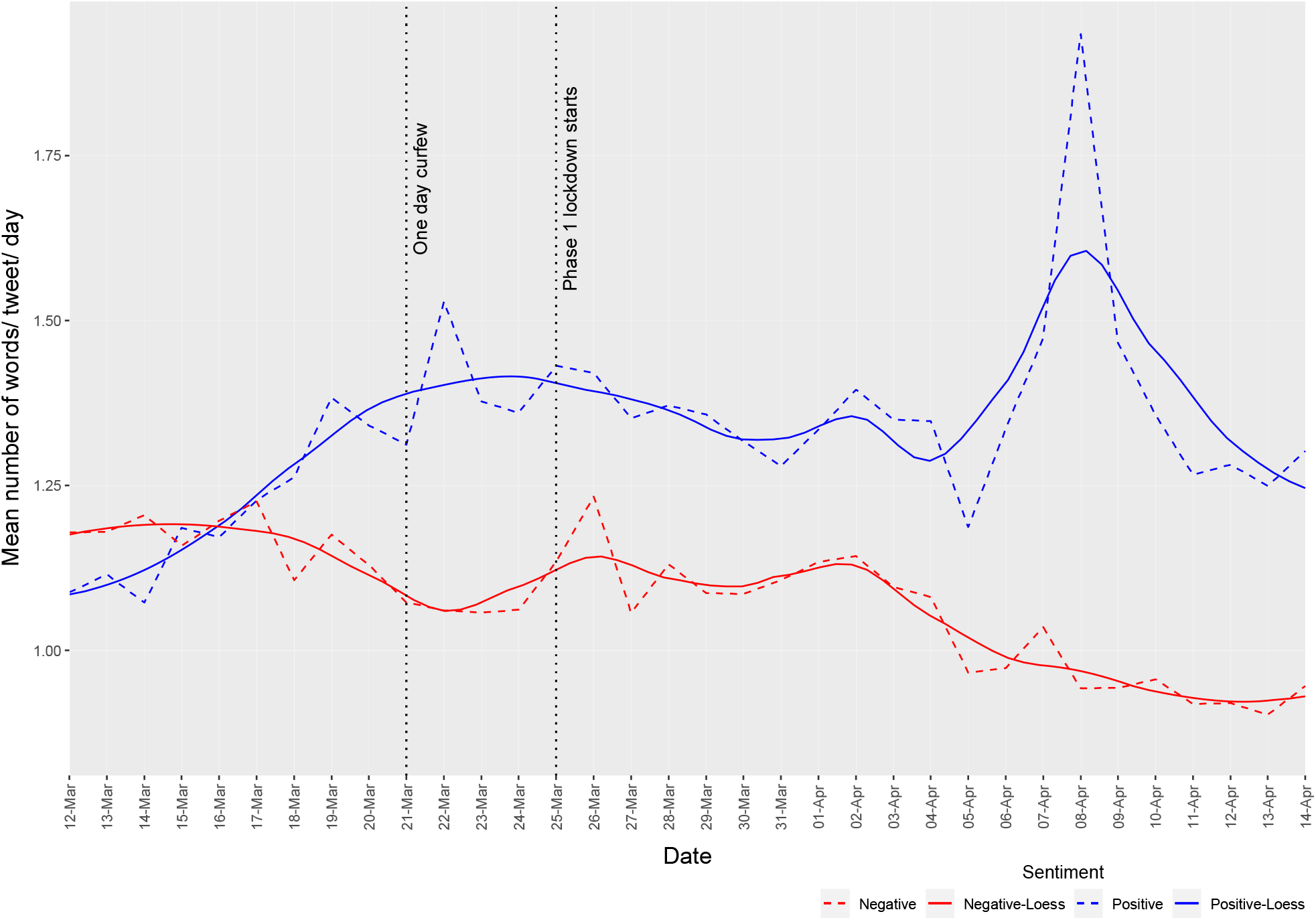
Trends in positive and negative public sentiments before (12.3.20 to 24.3.20) and during (25.3.20 to 14.4.20) the first phase of lockdown in India.

### 3.2. Emotions

The mean number of words per tweet representing the different emotions in descending order were as follows - trust (0.85), fear (0.66), anticipation (0.58), joy (0.46), sadness (0.40), anger (0.39), surprise (0.23), and disgust (0.20). There wasn’t much discernible change in the ranks of the emotions in terms of mean number of words per tweet or percentage of tweets with the emotion comparing the two time periods.(Table 1, Figure 3) Mean number of words per tweet reflecting trust was higher (0.84 words per tweet before and 0.85 words per tweet during the lockdown) than the mean number of words reflecting other emotions. Also, trust was the most common (52.6% before and 52.4% during the lockdown) emotion in the tweets followed by fear and anticipation.(Table 1)

**Figure 3.**
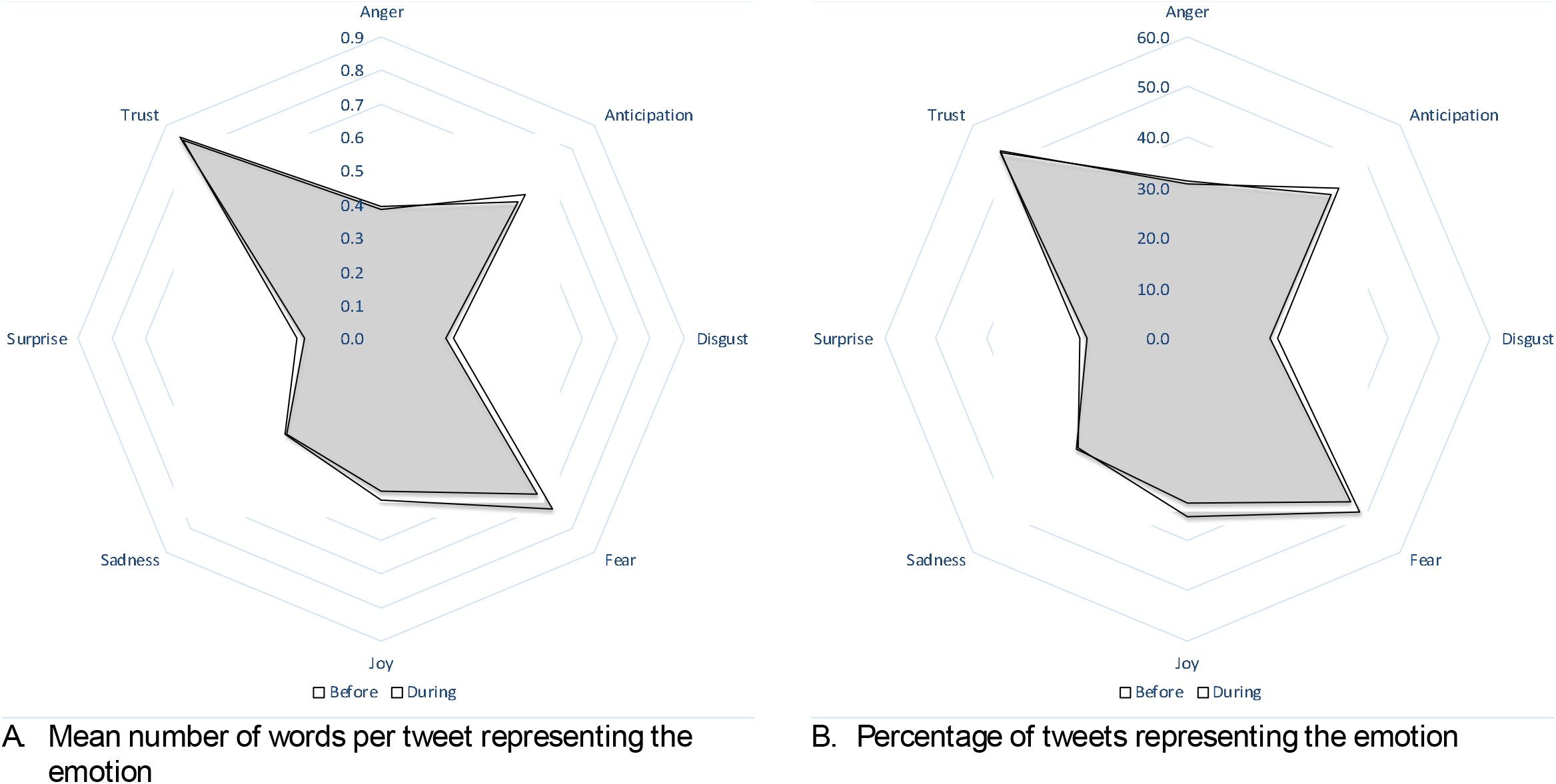
Pattern of public emotions before (12.3.20 to 24.3.20) and during (25.3.20 to 14.4.20) the first phase of lockdown in India.

### 3.3. City level analysis

The city-wise analysis of mean sentiment score and other emotions for the five cities namely Bhopal, Chennai, Delhi, Kolkata and Mumbai are presented are presented in Table 2 and Figure 4. In all the cities, the sentiment score shows a deliberate upward trend in the period after the lockdown announcement with Chennai showing more exuberance than the other cities, especially towards the end.(Figure 4)

**Table 2.** Public sentiments and emotions about COVID-19 in five major cities of India between March 12^th^ and April 14^th^, 2020.

| Place | Sentiment score, Mean (Range) | Number of words per tweet representing the sentiment, Mean (Range) |  | Number of words per tweet representing the emotion, Mean (Range) |  |  |  |  |  |  |  |
| --- | --- | --- | --- | --- | --- | --- | --- | --- | --- | --- | --- |
|  |  | Negative | Positive | Anger | Anticipation | Disgust | Fear | Joy | Sadness | Surprise | Trust |
| Delhi | 0.21 (16.5) | 0.99 (11) | 1.30 (12) | 0.40 (8) | 0.57 (9) | 0.19 (6) | 0.68 (9) | 0.43 (8) | 0.40 (8) | 0.23 (5) | 0.84 (9) |
| Mumbai | 0.23 (14.5) | 0.98 (9) | 1.32 (10) | 0.39 (6) | 0.57 (7) | 0.20 (6) | 0.66 (7) | 0.45 (8) | 0.39 (6) | 0.24 (4) | 0.85 (8) |
| Chennai | 0.37 (16.9) | 0.95 (11) | 1.52 (10) | 0.35 (7) | 0.62 (7) | 0.19 (5) | 0.61 (7) | 0.52 (8) | 0.41 (6) | 0.22 (5) | 0.83 (7) |
| Kolkata | 0.22 (14.7) | 1.01 (9) | 1.31 (13) | 0.41 (7) | 0.55 (8) | 0.20 (5) | 0.71 (7) | 0.44 (10) | 0.42 (6) | 0.23 (4) | 0.87 (10) |
| Bhopal | 0.23 (12.1) | 1.03 (9) | 1.35 (9) | 0.43 (6) | 0.58 (6) | 0.22 (5) | 0.70 (7) | 0.48 (6) | 0.39 (5) | 0.25 (4) | 0.88 (7) |

**Figure 4.**
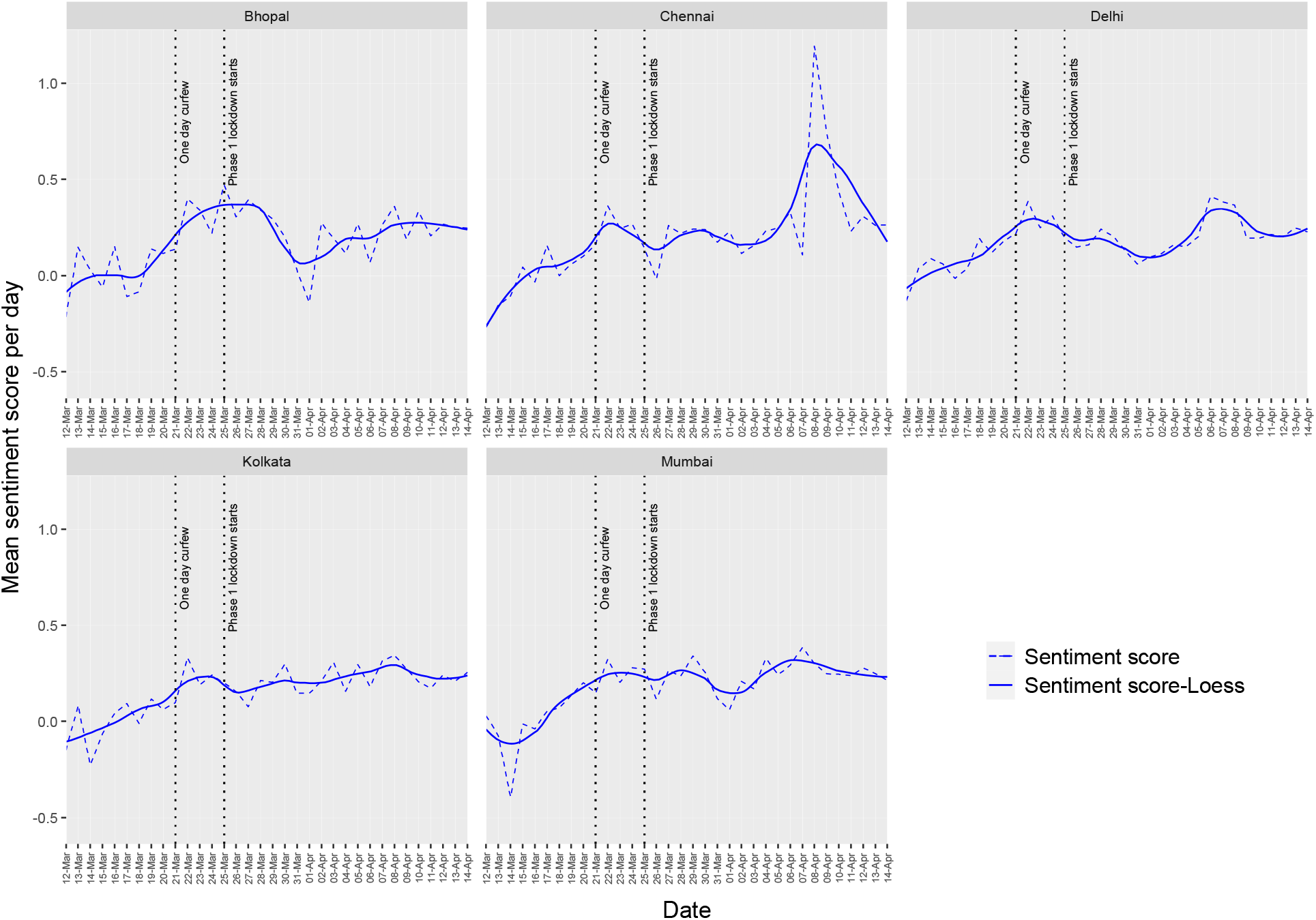
Trends in daily mean sentiment score before (12.3.20 to 24.3.20) and during (25.3.20 to 14.4.20) the first phase of lockdown in five major cities of India.

### 3.4. Change point analysis

There were three change points for sentiment score, the first two (on day -7 and 11) when a rise was noted and the third (on day 15) when it started to decrease. Positive sentiment mirrored the pattern of the sentiment score and negative sentiment followed a declining trend at similar changepoints. Trust continued to remain high after the first change point (on day - 8).(Table 3, Figure 5,6)

**Table 3.**
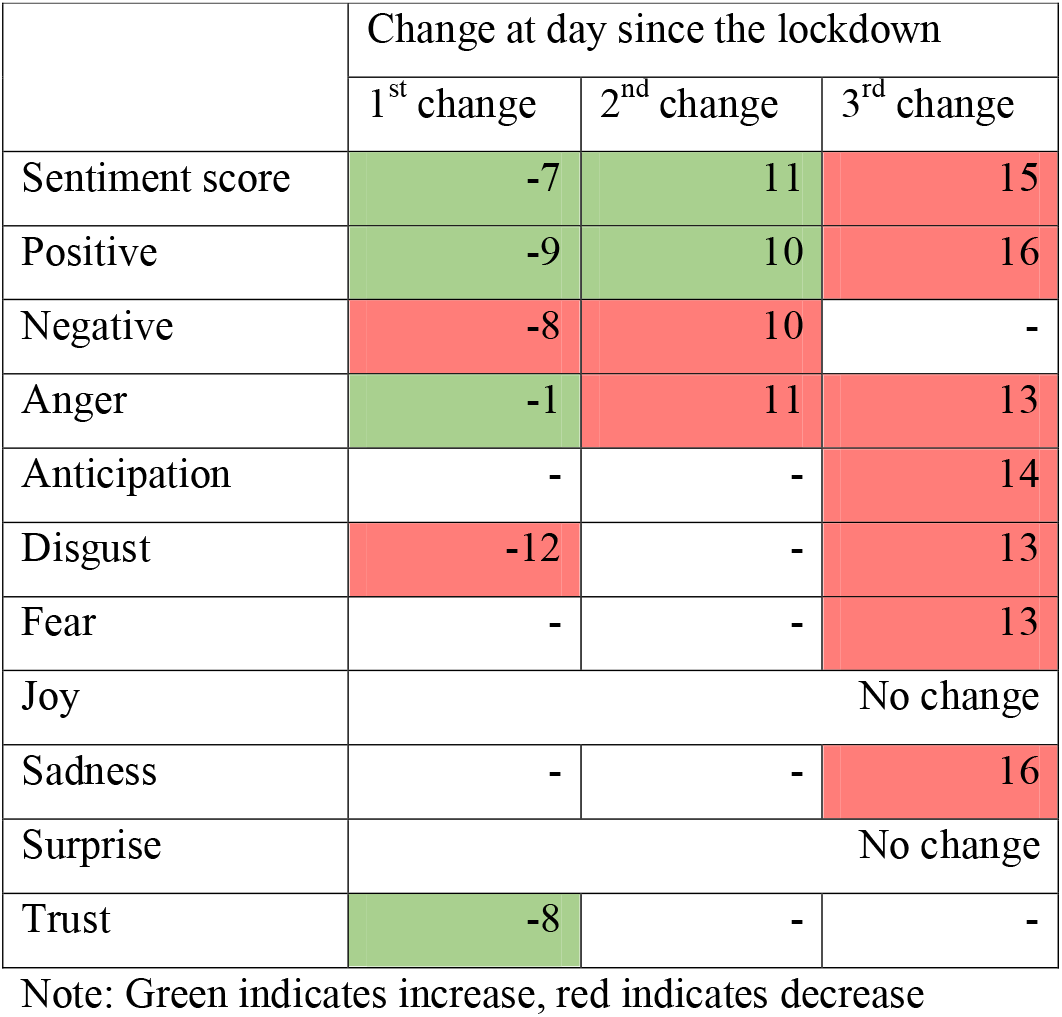
Change point analysis of the public sentiments and emotions in the first phase of lockdown in India between March 12th and April 14th, 2020.

|  | Change at day since the lockdown |  |  |
| --- | --- | --- | --- |
|  | 1<sup>st</sup> change | 2<sup>nd</sup> change | 3<sup>rd</sup> change |
| Sentiment score | -7 | 11 | 15 |
| Positive | -9 | 10 | 16 |
| Negative | -8 | 10 | - |
| Anger | -1 | 11 | 13 |
| Anticipation | - | - | 14 |
| Disgust | -12 | - | 13 |
| Fear | - | - | 13 |
| Joy | No change |  |  |
| Sadness | - | - | 16 |
| Surprise | No change |  |  |
| Trust | -8 | - | - |

Note: Green indicates increase, red indicates decrease

**Figure 5.**
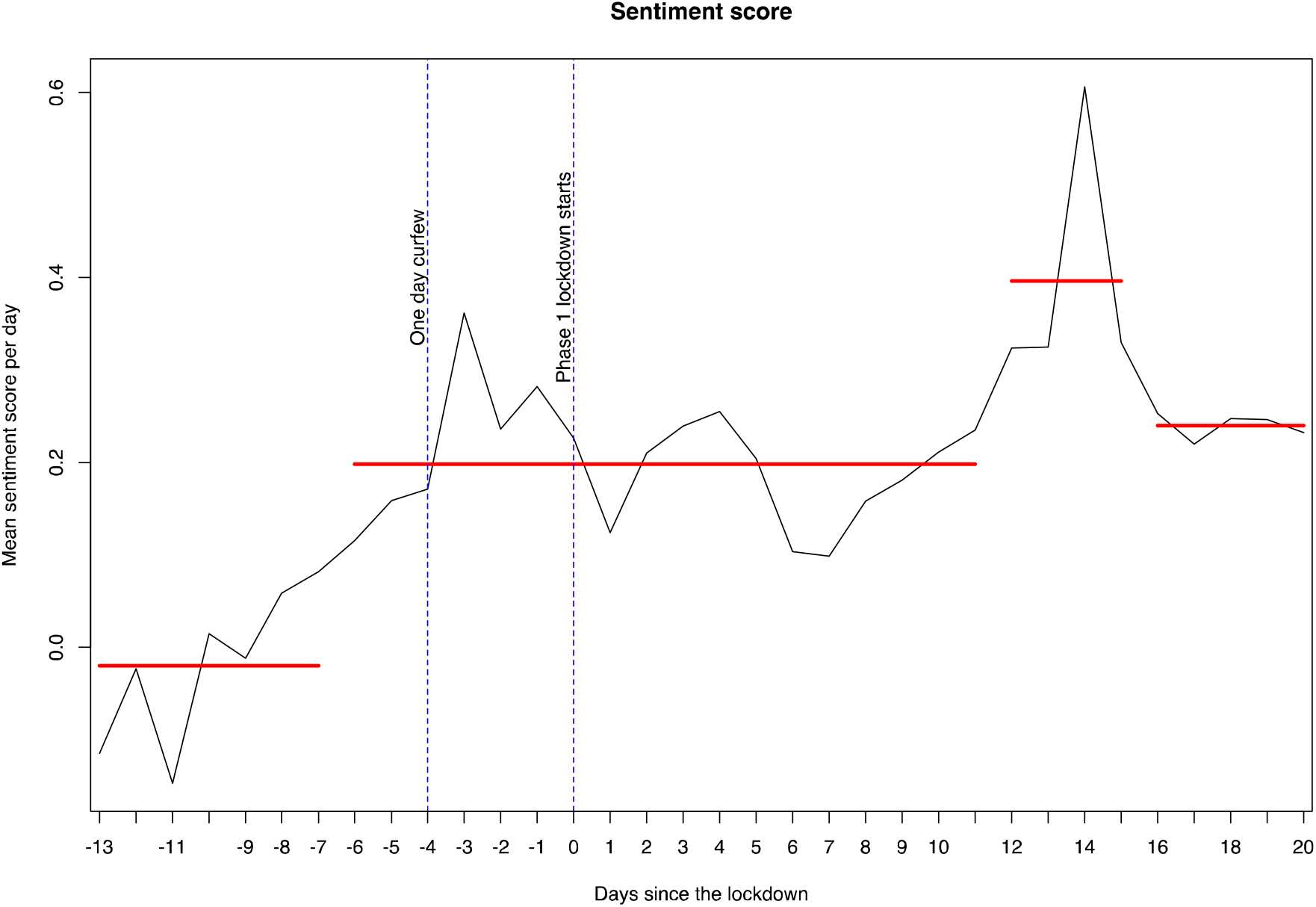
Change point analysis of the daily mean sentiment score in the first phase of lockdown in India between March 12th and April 14th, 2020.

**Figure 6.**
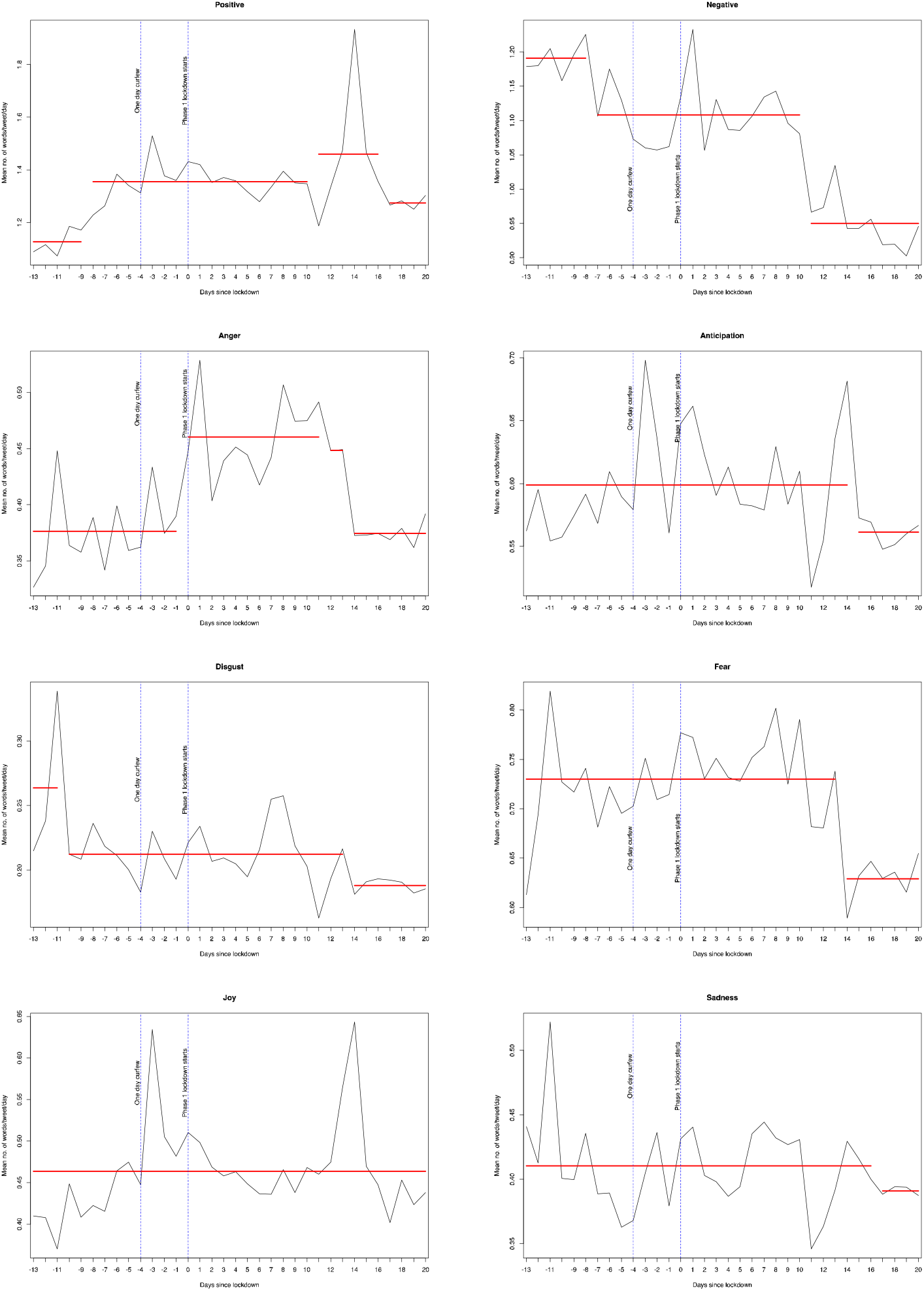
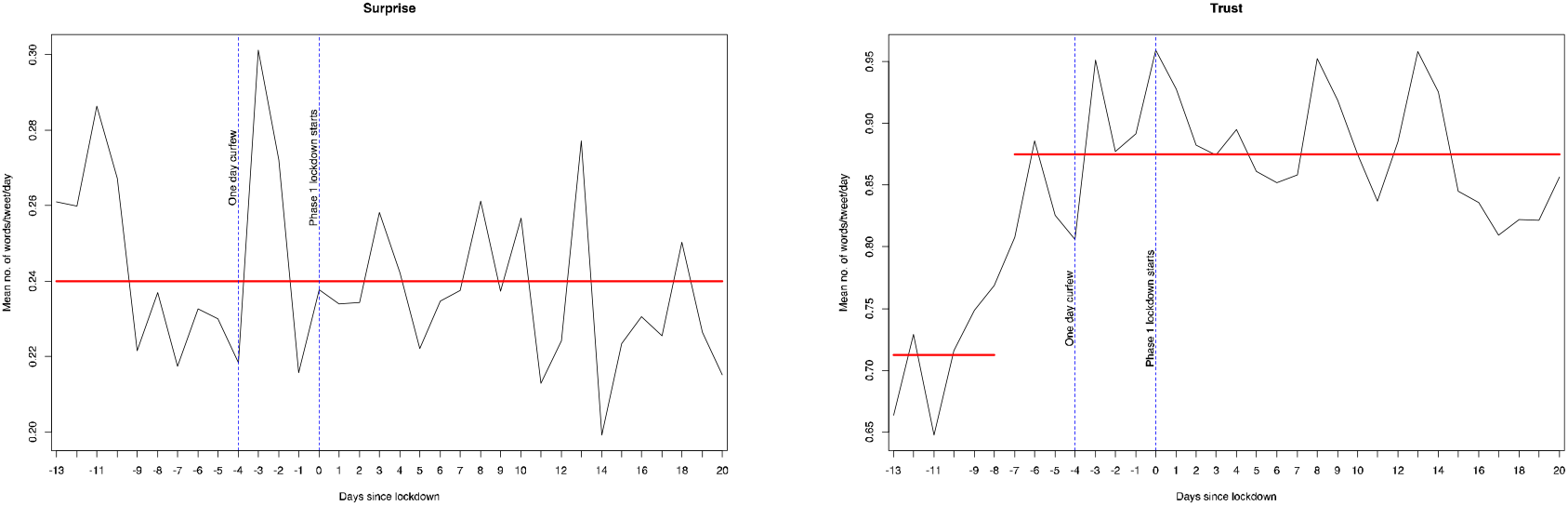
Change point analysis of public sentiments and emotions in the first phase of lockdown in India between March 12th and April 14th, 2020.

### 3.5. Word cloud

The word cloud portrays the 100 most commonly appearing words in the tweets during the study period.(Figure 7) As can be seen, before the lockdown the most prominent words used were ‘virus’, ‘curfew’, ‘face’ and ‘wash’ and during the lockdown the most prominent words were ‘lockdown’, ‘cases’, ‘warriors’ and ‘state’.

**Figure 7.**
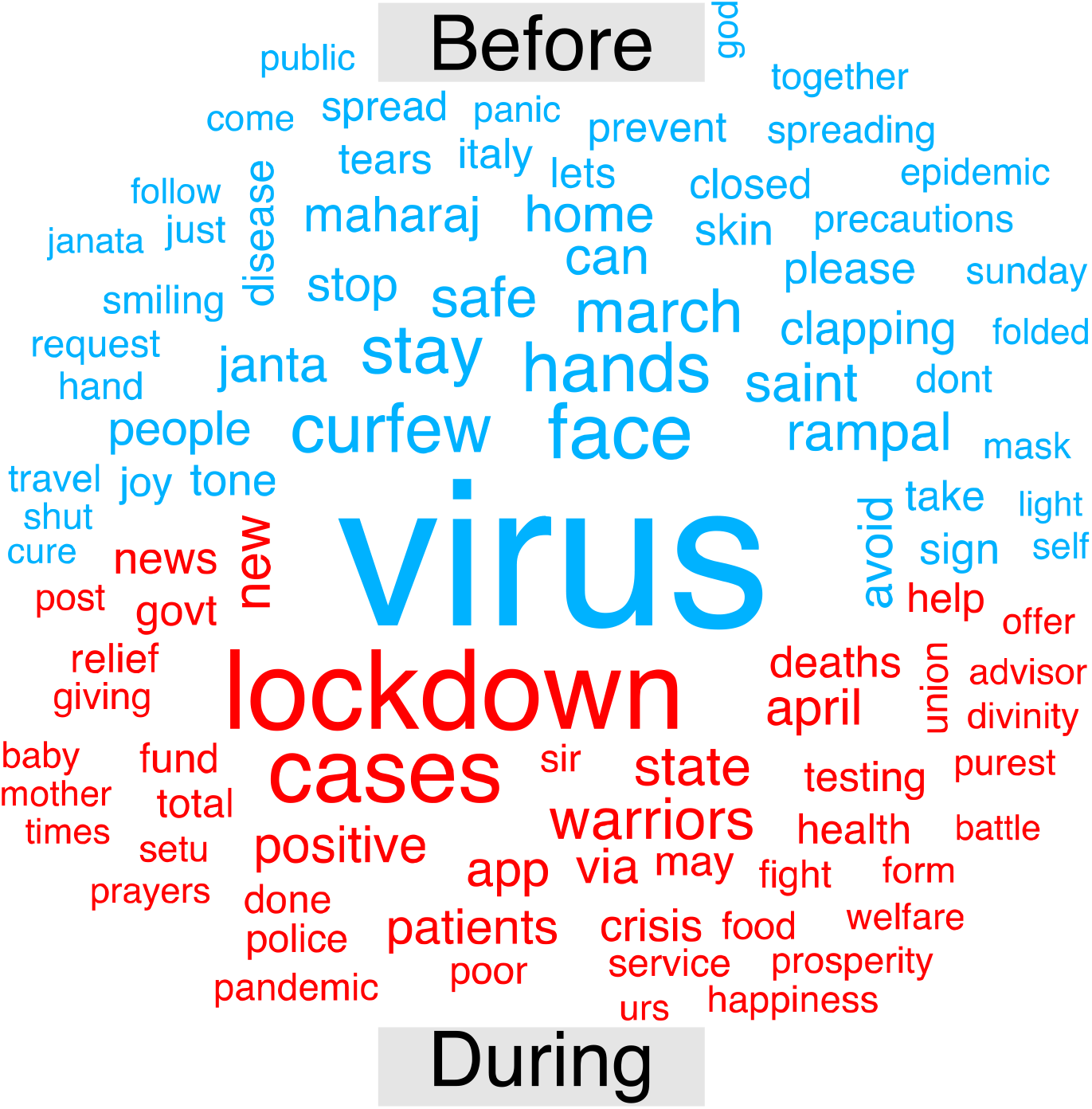
Word cloud reflecting public sentiment before (12.3.20 to 24.3.20) and during (25.3.20 to 14.4.20) the first phase of lockdown in India.

## 4. Discussion

### 4.1. Summary of main findings

Our sentiment analysis of 0.22 million tweets from five major cities in India between 12^th^ March 2020 to 14^th^ April 2020 reflected that the public sentiment was predominantly positive. Positive sentiments increased and negative sentiments decreased after the implementation of the lockdown. Trust and fear were the most common emotions expressed. Negative emotions namely anger, fear and sadness reduced during the lockdown. A positive public sentiment for the strategies implemented by the government in response to COVID-19 has implications for their compliance. Ultimately, this is expected to positively impact the course of the pandemic in the country.

### 4.2. Evidence from previous work

Studies capturing the sentiments about COVID-19 from China, the epicentre of the pandemic, report mixed results. Similar to our study, Zhao et al report their sentiment analysis of the microblogging site Sina Microblog. They observed that the emotional tendency of the public went from negative to neutral, with negative emotions weakening and positive emotions increasing.(Zhao et al. 2020) On the contrary, Li S et al in their analysis of sentiments of Weibo users before and after the declaration of COVID-19 outbreak report that negative emotions like anger increased and positive emotions decreased. People were most concerned about health and family while they were less concerned about leisure and friends. They emphasise the need for such analysis which fills the knowledge gaps of short-term individual changes in psychological conditions after a major event.(Li et al. 2020) Barkur et al in their letter to the editor mention that the emotion ‘trust’ stood out in India over all others. However, their analysis was very preliminary and considered data of only three days post the implementation of the lockdown.(Barkur, Vibha, and Kamath 2020)

### 4.3. Implications of the findings

The most prevalent emotion found in our analysis was trust. Public trust has been repeatedly shown in previous traditional surveys to be a prerequisite for the success of public health interventions such as a pandemic response.(Prati, Pietrantoni, and Zani 2011; Siegrist and Zingg 2014) Sociological models like the ‘trust-confidence-cooperation’ framework inform us that trust is what ultimately leads to public cooperation.(Earle and Siegrist 2008) In India, public trust seems to be in place and it is fertile grounds for the government to implement interventions for the larger public interest. It would be prudent of any government to engage in activities that keep the public trust high so that further public health actions would also be received and followed favourably. Fear was the most prevalent negative emotion that prevailed before and during the lockdown. This could be explained by the fact that this is an unprecedented situation the world finds itself in, in terms of the disease and the control measures. Fear is one of the most basic of human emotions. It is only natural and understandable that the public express fear as a major emotion in response to a novel, rapidly spreading disease and the possible adverse socioeconomic impact of the control measures.

The preponderance of positive sentiment and its increase post announcement of the lockdown could reflect an overall trust that the public have in the Government’s strategies to curb the pandemic. Another important reason for this increase in positivity could be explained by the popular characteristics of Twitter users. Twitters users are relatively more educated, younger, more likely to own a smartphone with Internet connection and work in the organised sector with a regular income.(Sloan et al. 2015, 2013) They are less likely to be have been affected economically and socially unlike the people working in the unorganised sector who faced wage losses and hunger due to the lockdown. These advantages that Twitter users enjoy might have influenced the trend towards the positivity.

The results of our study provide insights and valuable feedback to the policy makers. They can try to understand the pattern of negative emotions and work on addressing them through effective risk communication or adapting strategies to be more inclusive and suit local context. Packaging of risk communication and proposed interventions can either lead to a successful implementation or lead to rotting without public ownership.(Tavoschi et al. 2020) They can capitalise on the positive sentiments to draw lessons for future framing of public friendly strategies. It could also be vital to quickly pick up fake messages and dispel misinformation.(Porreca, Scozzari, and Di Nicola 2020)

Marketing agencies benefit greatly from their analysis of public sentiments to strategically and effectively widen their clientele.(Reyes-Menendez, Saura, and Filipe 2020) Public health, especially in low and middle income countries, is still in its infancy when it comes to monitoring and utilizing public emotions on social media for decision making. There is a need for building and strengthening surveillance systems for monitoring public sentiments on social media and use that knowledge to frame appropriate and responsive strategies. (Abd-Alrazaq et al. 2020) While the use of computer-based algorithms to draw inferences about human sentiments is promising, prediction by a computerized system alone is not holistic. A composite approach with a “human-in-the-loop” is recommended to provide the system with feedback. (Kunneman et al. 2020)

### 4.4. Strengths and limitations

To the best of our knowledge, this is the first comprehensive sentiment analysis of COVID-19 related tweets performed in India. By studying five major cities and areas around them we have covered a vast expanse of the country. We are also confident that the large number of tweets analysed along with the long duration over which they were captured provide a reasonably accurate and comprehensive assessment of the public sentiment about this major public health situation. Sentiment analysis is relatively new and multiple methods are adopted by different researchers not all of which have been tested on large datasets.(Gohil, Vuik, and Darzi 2018) Our study adopts a standard methodology for sentiment analysis and we provide a comprehensive description of the same enabling reproduction.

There are a few limitations to this study. We expect that Twitter users will be systematically different from non-users in terms of sociodemographic characteristics (age, gender, rurality, education, occupation, religious belief, political orientation and personality traits) which is a disadvantage shared by most social media-based sentiment analyses. However, sentiment analysis of social media data is the most practical way to reliably study real-time public sentiments when a pandemic situation like COVID-19 precludes the use of traditional research methods. In this study, we have used data from Twitter only and no other social media sites. Because, Twitter has been increasingly used by official agencies like ministries and public health organisations to send reports and communications to the common man, it is was judged more appropriate for the study objective. North-eastern states are not represented in our analysis because they reported <1% of the total number of cases in India.

### 4.5. Conclusion and recommendations

Positive sentiments dominated the COVID-19 outbreak in India even during the implementation of stringent control measures like complete national level lockdown. Whether this translated into optimal compliance with the interventions and effective control of COVID-19 in India is a subject for future research. Public health decision makers can use the understanding of these sentiments to guide their policy and risk communication strategies. In a rapidly evolving health crisis like COVID-19 where every day is different and challenging, a continuous monitoring of social media-based public sentiments can serve as an essential tool for decision making.

## Data Availability

All data produced in the present study are available upon reasonable request to the authors

## References

Abd-Alrazaq, Alaa, Dari Alhuwail, Mowafa Househ, Mounir Hamdi, and Zubair Shah. 2020. “Top Concerns of Tweeters During the COVID-19 Pandemic: Infoveillance Study.” Journal of Medical Internet Research 22 (4): e19016. https://doi.org/10.2196/19016.

Barkur, Gopalkrishna Vibha, and Giridhar B. Kamath. 2020. “Sentiment Analysis of Nationwide Lockdown Due to COVID 19 Outbreak: Evidence from India.” Asian Journal of Psychiatry 51 (June). https://doi.org/10.1016/j.ajp.2020.102089.

Braunack-Mayer, Annette J., Jackie M. Street, Wendy A. Rogers, Rodney Givney, John R. Moss, and Janet E. Hiller. 2010. “Including the Public in Pandemic Planning: A Deliberative Approach.” BMC Public Health 10 (1): 501. https://doi.org/10.1186/1471-2458-10-501.

Earle, T., and M. Siegrist. 2008. “Trust, Confidence and Cooperation Model: A Framework for Understanding the Relation between Trust and Risk Perception.” International Journal of Global Environmental Issues. https://doi.org/10.1504/IJGENVI.2008.017257.

Feinerer, Ingo. 2019. “Introduction to the Tm Package Text Mining in R.” 2019. https://cran.r-project.org/web/packages/tm/vignettes/tm.pdf.

Gabarron, Elia, Enrique Dorronzoro, Octavio Rivera-Romero, and Rolf Wynn. 2019. “Diabetes on Twitter: A Sentiment Analysis.” Journal of Diabetes Science and Technology 13 (3): 439–44. https://doi.org/10.1177/1932296818811679.

Gohil, Sunir, Sabine Vuik, and Ara Darzi. 2018. “Sentiment Analysis of Health Care Tweets: Review of the Methods Used.” Journal of Medical Internet Research. Journal of Medical Internet Research. https://doi.org/10.2196/publichealth.5789.

“Introduction to the Syuzhet Package.” n.d. Accessed May 12, 2020. https://cran.r-project.org/web/packages/syuzhet/vignettes/syuzhet-vignette.html.

Kang, Gloria J., Sinclair R. Ewing-Nelson, Lauren Mackey, James T. Schlitt, Achla Marathe, Kaja M. Abbas, and Samarth Swarup. 2017. “Semantic Network Analysis of Vaccine Sentiment in Online Social Media.” Vaccine 35 (29): 3621–38. https://doi.org/10.1016/j.vaccine.2017.05.052.

Kim, Hye Kyung, and Jeff Niederdeppe. 2013. “The Role of Emotional Response during an H1N1 Influenza Pandemic on a College Campus.” Journal of Public Relations Research 25 (1): 30–50. https://doi.org/10.1080/1062726X.2013.739100.

Kumar, Akshi, and Teeja Mary Sebastian. 2012. “Sentiment Analysis on Twitter.” International Journal of Computer Science Issues 9 (4).

Kunneman, Florian, Mattijs Lambooij, Albert Wong, Antal Van Den Bosch, and Liesbeth Mollema. 2020. “Monitoring Stance towards Vaccination in Twitter Messages.” BMC Medical Informatics and Decision Making 20 (1). https://doi.org/10.1186/s12911-020-1046-y.

Li, Sijia, Yilin Wang, Jia Xue, Nan Zhao, and Tingshao Zhu. 2020. “The Impact of Covid-19 Epidemic Declaration on Psychological Consequences: A Study on Active Weibo Users.” International Journal of Environmental Research and Public Health 17 (6). https://doi.org/10.3390/ijerph17062032.

Liu, Qian, Zequan Zheng, Jiabin Zheng, Qiuyi Chen, Guan Liu, Sihan Chen, Bojia Chu, et al. 2020. “Health Communication Through News Media During the Early Stage of the COVID-19 Outbreak in China: A Digital Topic Modeling Approach (Preprint).” Journal of Medical Internet Research, April. https://doi.org/10.2196/19118.

Loft, Louise H., Eva A. Pedersen, Stine U. Jacobsen, Bolette Søborg, and Janne Bigaard. 2020. “Using Facebook to Increase Coverage of HPV Vaccination among Danish Girls: An Assessment of a Danish Social Media Campaign.” Vaccine. https://doi.org/10.1016/j.vaccine.2020.04.032.

Ministry of Home Affairs. n.d. Order No. 40-3/2020-DM-I(A). New Delhi India.

“NRC Emotion Lexicon.” n.d. Accessed May 12, 2020. https://saifmohammad.com/WebPages/NRC-Emotion-Lexicon.htm.

Phani, Shanta, Shibamouli Lahiri, and Arindam Biswas. 2016. “Sentiment Analysis of Tweets in Three Indian Languages.” In Proceedings of the 6th Workshop on South and Southeast Asian Natural Language Processing, 93–102. Osaka.

Porreca, Annamaria, Francesca Scozzari, and Marta Di Nicola. 2020. “Using Text Mining and Sentiment Analysis to Analyse YouTube Italian Videos Concerning Vaccination.” BMC Public Health 20 (1). https://doi.org/10.1186/s12889-020-8342-4.

Prati, Gabriele, Luca Pietrantoni, and Bruna Zani. 2011. “Compliance with Recommendations for Pandemic Influenza H1N1 2009: The Role of Trust and Personal Beliefs.” Health Education Research 26 (5): 761–69. https://doi.org/10.1093/her/cyr035.

Reintjes, Ralf, Enny Das, Celine Klemm, Jan Hendrik Richardus, Verena Keßler, and Amena Ahmad. 2016. “‘Pandemic Public Health Paradox’: Time Series Analysis of the 2009/10 Influenza A/H1N1 Epidemiology, Media Attention, Risk Perception and Public Reactions in 5 European Countries.” PLoS ONE 11 (3). https://doi.org/10.1371/journal.pone.0151258.

Reyes-Menendez, Ana, Jose Ramon Saura, and Ferrão Filipe. 2020. “Marketing Challenges in the #MeToo Era: Gaining Business Insights Using an Exploratory Sentiment Analysis.” Heliyon 6 (3). https://doi.org/10.1016/j.heliyon.2020.e03626.

Siegrist, Michael, and Alexandra Zingg. 2014. “The Role of Public Trust During Pandemics.” http://Dx.Doi.Org/10.1027/1016-9040/A000169, January. https://doi.org/10.1027/1016-9040/A000169.

Sloan, Luke, Jeffrey Morgan, Pete Burnap, and Matthew Williams. 2015. “Who Tweets? Deriving the Demographic Characteristics of Age, Occupation and Social Class from Twitter User Meta-Data.” https://doi.org/10.1371/journal.pone.0115545.

Sloan, Luke, Jeffrey Morgan, William Housley, Matthew Williams, Adam Edwards, Pete Burnap, and Omer Rana. 2013. “Knowing the Tweeters: Deriving Sociologically Relevant Demographics from Twitter.” Sociological Research Online 18 (3): 74–84. https://doi.org/10.5153/sro.3001.

Tavoschi, Lara, Filippo Quattrone, Eleonora D’Andrea, Pietro Ducange, Marco Vabanesi, Francesco Marcelloni, and Pier Luigi Lopalco. 2020. “Twitter as a Sentinel Tool to Monitor Public Opinion on Vaccination: An Opinion Mining Analysis from September 2016 to August 2017 in Italy.” Human Vaccines and Immunotherapeutics. https://doi.org/10.1080/21645515.2020.1714311.

“Twitter Terms of Service.” n.d. Accessed May 12, 2020. https://twitter.com/en/tos.

Zhao, Yuxin, Sixiang Cheng, Xiaoyan Yu, and Huilan Xu. 2020. “Chinese Public Attention to COVID-19 Epidemic: Based on Social Media (Preprint).” Journal of Medical Internet Research 22 (5). https://doi.org/10.2196/18825.

